# Impact Assessment of Full and Partial Stay-at-Home Orders, Face Mask Usage, and Contact Tracing: An Agent-Based Simulation Study of COVID-19 for an Urban Region

**DOI:** 10.1101/2020.07.27.20163121

**Authors:** Hanisha Tatapudi, Rachita Das, Tapas K. Das

**Affiliations:** Department of Industrial and Management System Engineering, University of South Florida, Tampa, Florida, USA; Miller School of Medicine, University of Miami, Miami, Florida, USA

**Keywords:** COVID-19, SARS-CoV-2, Agent-based simulation model, mitigation strategies, face mask, contact tracing

## Abstract

**Purpose:** Various social intervention strategies to mitigate COVID-19 are examined using a comprehensive agent-based simulation model. A case study is conducted using a large urban region, Miami-Dade County, Florida, USA. Results are intended to serve as a planning guide for public health decision makers.

**Methods:** The simulation model mimics daily social mixing behavior of the susceptible and infected generating the spread. Data representing demographics of the region, virus epidemiology, and social interventions shapes model behavior. Results include daily values of infected, reported, hospitalized, and dead.

**Results:** Study results show that stay-at-home order is quite effective in flattening and then reversing the case growth curve subsiding the pandemic with only 5.8% of the population infected. Whereas, following Florida’s current Phase II reopening plan could end the pandemic via herd immunity with 75% people infected. Use of surgical variety face masks reduced infected by 20%. A further reduction of 66% was achieved through contact tracing.

**Conclusions:** For Miami-Dade County, a strategy comprising mandatory use of face masks and aggressive contact tracing to identify 50% of the asymptomatic and pre-symptomatic, if adopted now, can potentially steer the COVID-19 pandemic to subside within next 3 months with approximately one fifth of the population infected.

## INTRODUCTION

Emergence of the severe acute respiratory syndrome coronavirus type 2 (SARS-CoV-2) was first reported on December 31, 2019 in Wuhan, China and subsequently declared a global pandemic on March 11 by the World Health Organization (WHO) [1, 2]. As of July 22, 2020, the number of reported cases worldwide has reached over 14.9 million causing 617,730 deaths. The number of infected cases continues to rise quite significantly [3]. The U.S. has been among the hardest hit by the coronavirus pandemic with nearly 4 million reported infections and 142,500 reported deaths (∼23% of the total reported deaths worldwide) by July 22, 2020. However, as the new cases, hospital admissions, and deaths began to decline in mid-May, most States in the U.S. began phased lifting of their social intervention measures. For example, Florida adopted a three phased approach: Phase I (which began in May 18, 2020) allowed most business and workplaces to reopen with up to 50% of their building capacities and with large events constrained to 25%; Phase II began in June 5, 2020 and allowed all businesses to reopen for up to 50-75% of their capacities and also permitting events in large venues with no more than 50% of their capacities; Phase III will be akin to a complete reopening for which neither a date nor the criteria have been declared. As the reopening entered Phase II, Florida, along with many other states, began to see sharp increases in daily new infections (e.g., Florida reported over 15,000 new cases on July 11, 2020 along with a test positivity rate reaching over 15%).

In this paper, we investigate a few ‘what-if’ scenarios including if the stay-at-home order were not lifted, if the Phase II order continues unaltered, what impact will the mandatory face mask usage have on the infections and deaths, and finally, how do the benefits of contact tracing vary with tracing target levels.

## METHODOLOGY

Our research methodology involves development and use of a comprehensive agent-based (AB) simulation model for COVID-19. The model mimics hour by hour social mixing behavior at home, school, work, and community places for millions of people in a region of outbreak. The model uses detailed census reported demographics of age, household, workplaces, etc., virus epidemiology parameters, and the social interventions that are in place. The initial infected cases introduced to the simulation are those with travel histories to high risk regions/countries, and the mixing with these cases generates the community spread. The infected cases follow the SARS-CoV-2 disease natural history. The model output comprises the daily values of infected, reported, hospitalized, and dead. We calibrate the model using parameters that control the extent of virus sharing (transmission coefficient) and social mixing behavior in various phases of intervention orders. The model is validated by closely reproducing the numbers of reported cases and deaths in an urban region that is used as a case study; Miami-Dade County, Florida, USA with 2.8 million people. For a detailed description of the simulation model refer to the *supplementary document*. The supplementary document also includes our model calibration approach, validation results, and the Tables listing the data used in the simulation model.

## RESULTS

We used our model to predict the rate of growth of infected cases, reported cases, hospitalizations, and deaths for the case study region under various social intervention scenarios. First, we allowed the model to mimic retrospectively the progress of the pandemic under three separate intervention scenarios for a large number of days. The scenarios are: stay-at-home order continued without reopening until pandemic subsides, Phase I of reopening continued without moving into Phase II, and Phase II of reopening continued without the use of face mask or any other changes. Thereafter, we conducted a prospective examination of the impact we are likely to see in coming days from the use of face masks and contact tracing. A summary of some of our model results from the case study is presented in Table 1.

**Table 1.** Summary of the key results from the AB simulation model for a sample urban outbreak region (Miami-Dade County of Florida, USA) with population of 2.8 million

| Interventions →<br>Outcomes ↓ | If Stay-At-Home order were not lifted (started March 17, 2020) | If Phase I reopening continued (started May 18, 2020) | If Phase II reopening continues without alterations (started June 5, 2020) | If Phase II reopening continues with mandatory use of face masks (started June 25, 2020) | If Phase II reopening continues with use of face masks and contact tracing with 50% target (starting June 30, 2020) |
| --- | --- | --- | --- | --- | --- |
| Time when pandemic subsides below a threshold | Early Aug. 2020 | July **2021** | End-Oct. 2020 | End-Nov. 2020 | End-Sept. 2020 |
| Total number of infections (95% C.I.) | 162K<br>(136K – 188K) | 600K<br>(530K – 670K) | 2.17 million<br>(2.16 million – 2.18 million) | 1.74 million<br>(1.73 million – 1.75 million) | 581K<br>(447K – 716K) |
| Total number of reported cases (95% C.I.) | 23K<br>(19K – 27K) | 220K<br>(186K – 254K) | 866K<br>(854K – 877K) | 714K<br>(702K – 726K) | 247K<br>(178K – 316K) |
| Total number of hospitalizations (95% C.I.) | 4.1K<br>(3.5K – 4.8K) | 37.5K<br>(31.7K – 43.4K) | 149K<br>(147K – 151K) | 120K<br>(119K – 122K) | 35.2K<br>(25.6K – 44.8K) |
| Total number of deaths (95% C.I.) | 1K<br>(0.9K – 1.2K) | 9.4K<br>(7.9K – 10.8K) | 36.4K<br>(35.8K – 36.9K) | 29.7K<br>(29.3K – 30.2K) | 8.8K<br>(6.5K – 11.1K) |

In what follows, we discuss results from our case study. Figures 1 and 2 show the simulation results for the retrospective examination scenarios with average values (with 95% CIs in shade) of daily cumulative cases of actual infected, reported, hospitalized, and dead. The blue dotted lines represent the actual numbers of infected and dead as reported in the Florida COVID-19 dashboard till June 24 (our calibration period was till June 17). Figure 1 shows a strong influence of continuing with the stay-at-home order in curbing the COVID-19 growth within approximately 6 months from its inception with on average less than 5.8% of the population infected, 0.15% hospitalized, and 0.037% dead; 50 or below daily new infections was used as the criterion to consider that pandemic has subsided in Miami-Dade County.

**Figure 1.**
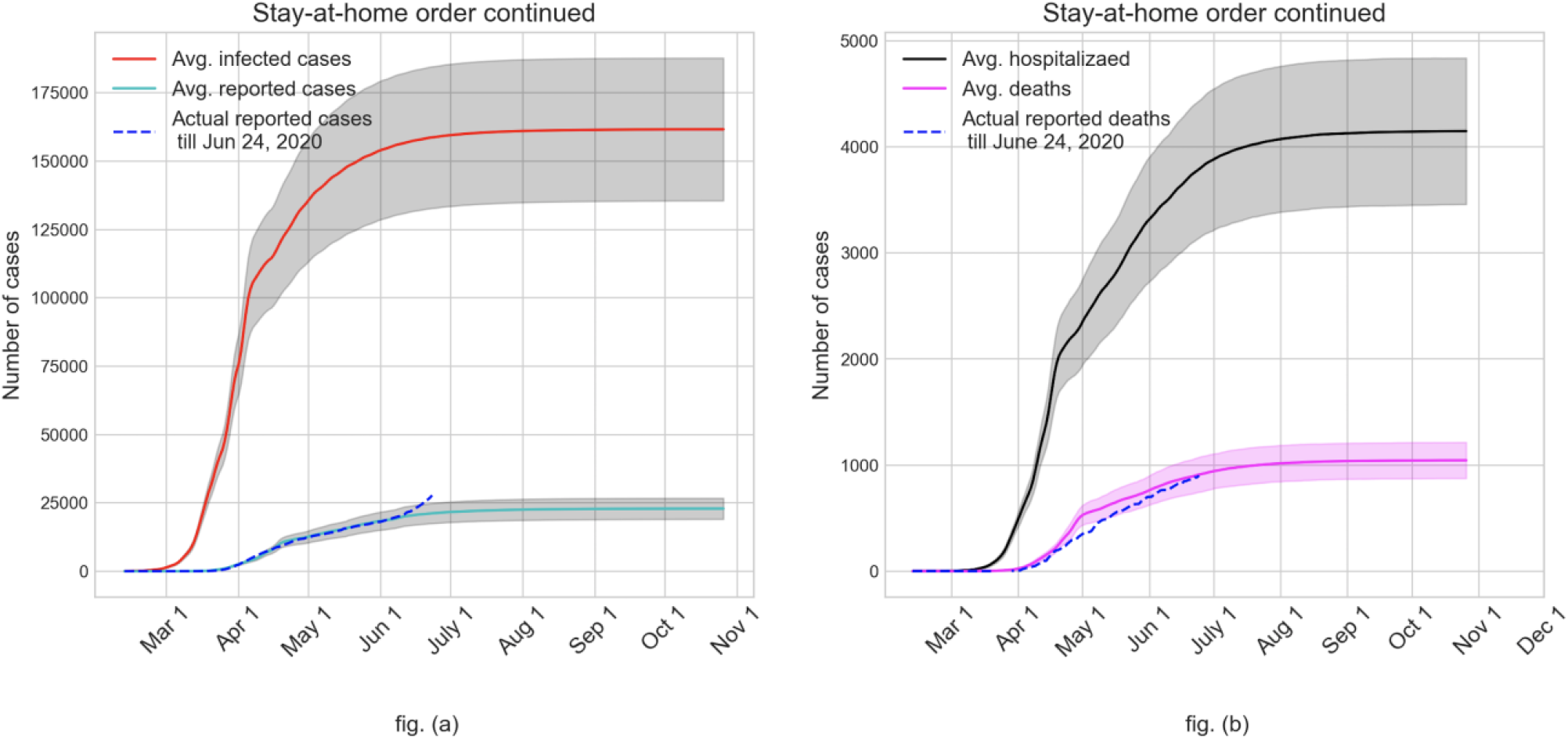
Growth of actual and reported infected cases (fig. a) and hospitalizations and deaths (fig. b) if stay-at-home order were not lifted

**Figure 2.**
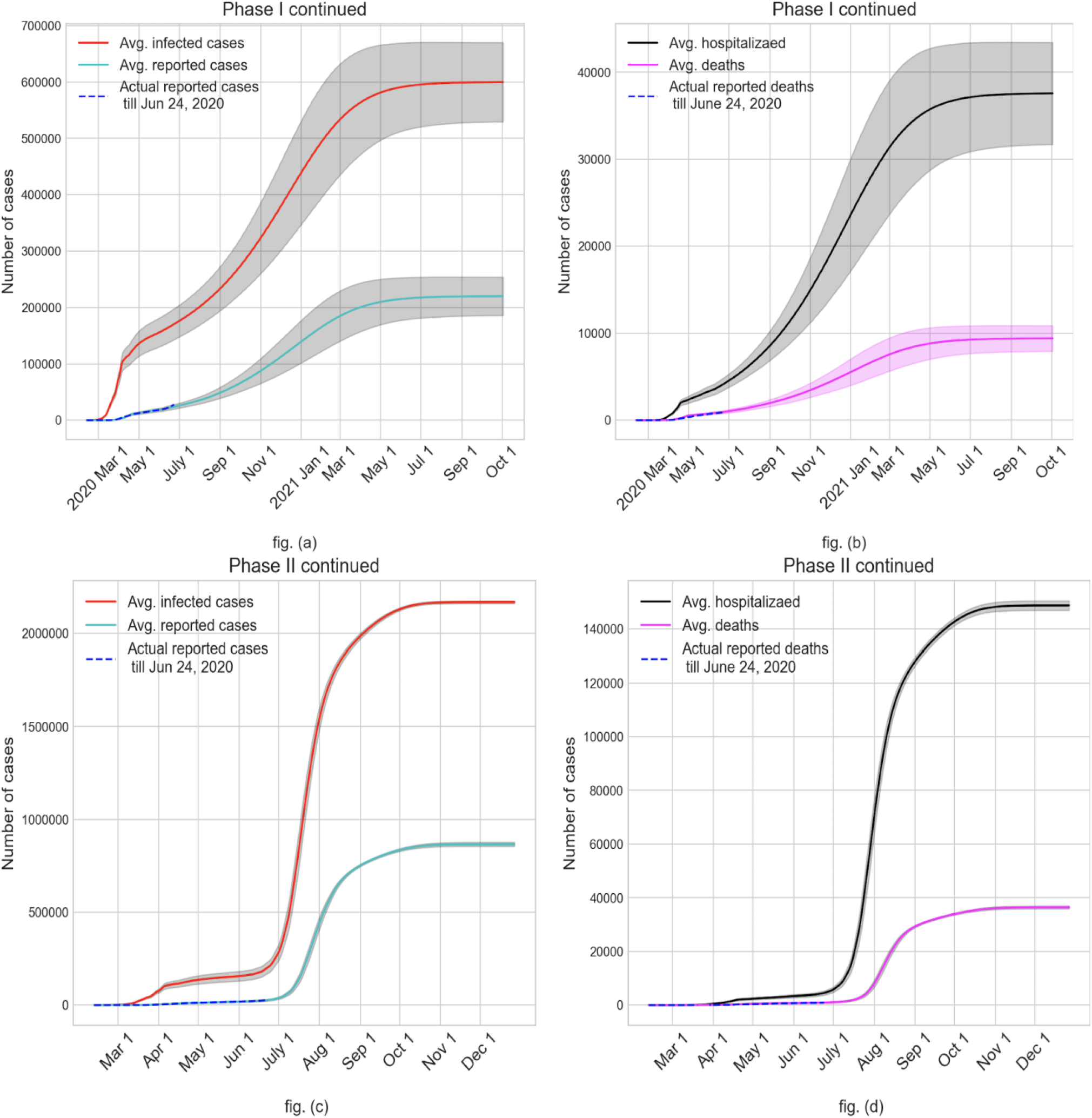
Case study outcomes (average values with 95% C.I.) of continuing with Phase I reopening (fig. a and fig. b) and Phase II reopening without face mask and contact tracing (fig. c and fig. d)

Figure 2 shows the expected outcomes of continuing with the Phase I order and the Phase II order. Figure 2(a) demonstrates a clear upward swing of the number of infected by the end of May as a result of Phase I reopening, in contrast to stay-at-home scenario where the numbers actually begin to drop at the end of May. The upward trajectory continues for nearly 12 months after reopening before curving down and subsiding the pandemic in July 2021. This scenario would have resulted in on average about 21% of the population infected (see fig. a), 1.3% hospitalized, and 0.34% dead (see fig. b). Figures 2(c) and 2(d) depict the rather grim outcome of continuing with Phase II order without face mask where over 75% of the population gets infected, 5.5% of the population hospitalized, and 1.3% dead. The steep multi-fold increase in the number of infected in late June after the Phase II opening in June 5 results in an end of the pandemic via herd immunity by late October 2020.

Hereafter, we used our model in a prospective examination of the pandemic progression under Phase II with the use of face mask and contact tracing. Mandatory use of face mask in work and community places where maintaining social distancing is not feasible was added to the Phase II guidelines starting June 25, 2020 in Miami-Dade County. In a recent article that analyzed data from the literature for SARS, MERS, and COVID-19 outbreaks, it is shown that adjusted odds ratio (aOR) of getting an infection after wearing surgical variety masks versus without wearing mask is 0.33 on average [4]. This can be interpreted as the likelihood of getting infected if wearing a surgical mask is one third of what it would be if not wearing a mask. Hence, we considered a 67% reduction in the transmission coefficient 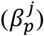 used in calculating the force of infection (see equation (1) in supplementary document), assuming a 100% compliance in the use of surgical variety masks at work and community places. We also tested the impact of 30% and 45% reductions in the transmission coefficient value, which translate to approximately 50% and 70% compliance for face mask usage, respectively. The anticipated impact of face mask usage together with Phase II order on the average cumulative numbers of infected are shown in Figure 3(a). It also depicts the risk difference between the average values of cumulative infected without and with the use of face mask considering a 100% compliance. It may be noted that since the infections grow slower with the use of face mask, the cumulative risk difference rises to almost 875K in the middle of August and then settles down close to 430K when pandemic subsides by the end of November, 2020. Figure 3(b) depicts the daily values of the average infected for Phase II without and with face mask for a 67% reduction in transmission coefficient (100% compliance). As expected, the peak of daily infection with face mask usage is shifted to a slightly later date and the downward trend begins after a smaller percentage (31%) of the total population being infected compared to 36% without the use of face mask.

**Figure 3.**
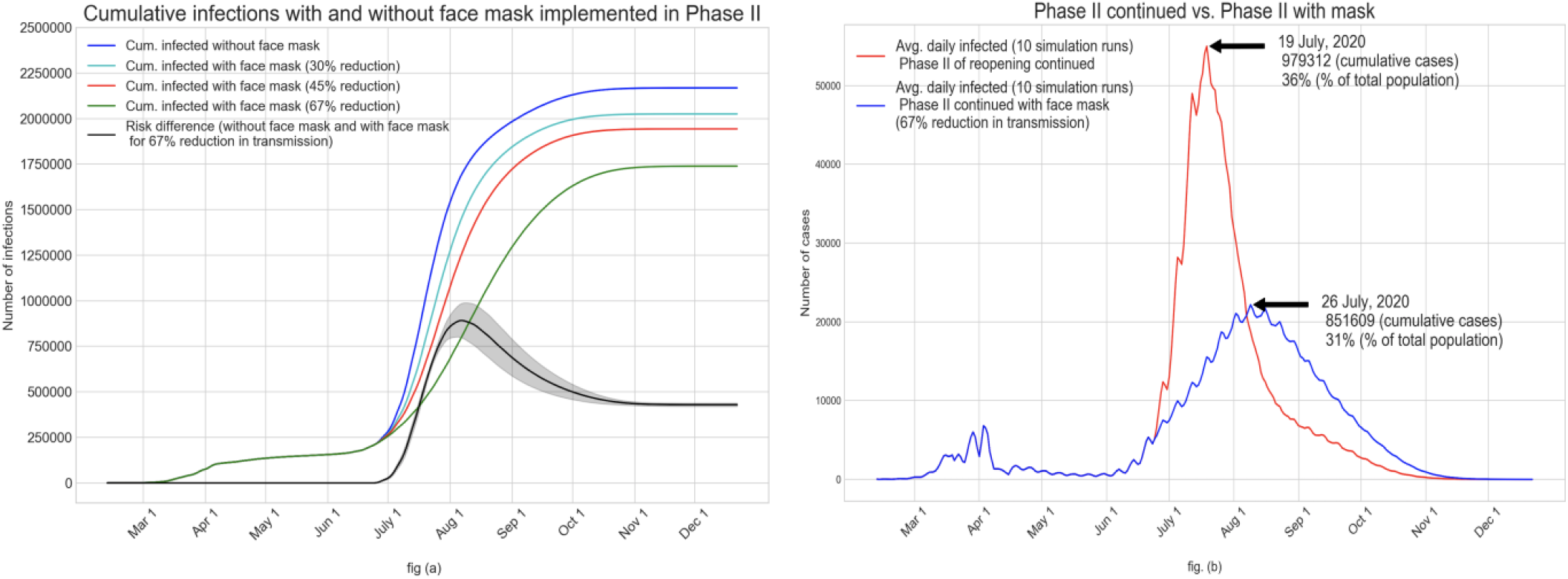
(fig. a) Impact of face mask usage starting June 25 (together with Phase II order) on the average cumulative infected for all compliance levels; (fig. b) Impact of 100% compliance of mask usage on the average daily infected

Though the use of mask with a 100% compliance together with the Phase II order is likely to reduce a large number of infections (an estimated 430K), this strategy still leaves a high percentage (63%) of total population infected before the pandemic subsides likely via reaching herd immunity. While a vaccine is still unavailable, it is likely that the only other way to reduce the size of this impacted population is to implement contact tracing. We used our model to examine a number of different contact tracing strategies by adding them to the scenario of Phase II with 100% face mask usage. We implemented contact tracing starting June 30 with a number of different targets (20%, 30%, 40%, and 50%) of identifying asymptomatic and pre-symptomatic cases. The impact on the average cumulative values of actual infected and the reported cases are shown in Figure 4(a). It can be observed that contact tracing can significantly reduce the number of people infected. With the 50% target for contact tracing (which is an aggressive goal), the average cumulative number of infected by the time the new infections fall below a threshold (possibly by the end of September, 2020) would reduce to 581K from over 1.73 million with Phase II and face mask alone (a 66% reduction). The corresponding reductions in cumulative infections and the associated times for pandemic to subside that can be expected from contact tracing targets of 40%, 30%, and 20% are 58% (mid-October), 41% (mid-November), and 14% (mid-December). It may also be noted that the impact of contact tracing target on the reduction of cumulative infected is nonlinear. Figure 4(b) shows the average cumulative numbers of hospitalizations and deaths. Expected reductions in hospitalization achieved from contact tracing targets of 50%, 40%, 30%, and 20% compared to the use of face mask alone (during Phase II) are 71%, 62%, 43%, and 14%, respectively. The corresponding expected reductions in the number of deaths are 70%, 62%, 43%, and 14%, respectively.

**Figure 4.**
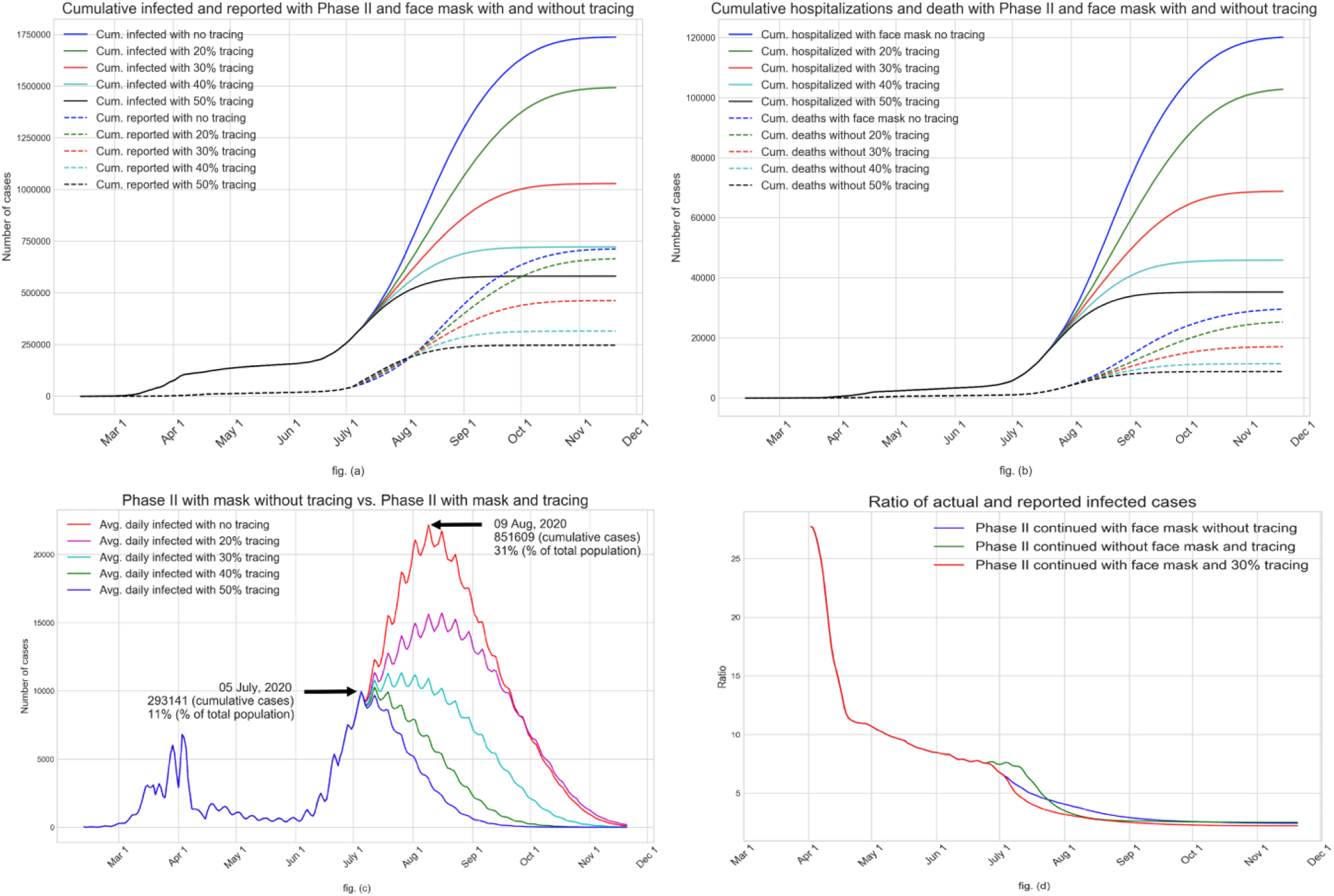
Impact of contact tracing, starting on June 30, during Phase II with face mask usage with 100% compliance

Figure 4(c) shows the impact of contact tracing starting on June 30, 2020 on the average daily infected values. It is interesting to note from the figure that an aggressive contact tracing (with associated testing and isolation of those found infected) appears to be capable of quickly turning the tide on new infections. Various COVID-19 dashboards maintained by government and private agencies have been reporting data including numbers of infected (tested positive), hospitalized, and dead. But the actual numbers of infected people in the outbreak regions remain a subject of expert opinion. Speculations abound place the ratio of actual to reported numbers of infected to as high as 10. As our simulation model yields estimates of the actual number of infected, we calculated the daily values of the ratio of average actual infected to average reported for a few scenarios: Phase II continued, Phase II with 100% face mask usage, and Phase II with face mask and contact tracing with 30% target. Values of these ratios are shown in Figure 4(d). It can be seen that in the initial days of the pandemic the ratios are very high (close to 30), which we believe is due to under testing together with long reporting delay. However, as the testing of the symptomatic increases and reporting delay decreases over time, the ratios come down sharply to 10 and continue to fall to near 7. The ratios further decreases gradually to about 2.5 as the daily new infections begin to fall in late July (see fig. 3b) and early August (see fig. 4c). As of mid-July, with surging numbers of daily new cases (see fig. 4c, Phase II with face mask), test reporting delay appears to have gone up to a week or more. We did not consider that in our simulation experiments. We note however that increased test reporting delay will reduce the beneficial impact of contact tracing.

## IV. DISCUSSION

We have developed an agent-based simulation model for COVID-19 pandemic to serve as a policy evaluation tool for public health decision makers. Similar AB models have been presented to the literature for simulating anticipated avian influenza pandemic outbreaks [5-10], to cite a few. Our simulation model is written in C/C++ and implemented using GNU General Public License [11].

Our model offers the flexibility to implement a variety of societal conditions including test availability, test reporting delay, stay-at-home order, partial reopening, selective closures of schools and workplaces when infections reappear, use of face mask with various levels of compliance, contact tracing, vaccinations, and use of antivirals. Only a subset of these conditions have been examined and reported in this paper. As we were completing this manuscript, the Miami-Dade County has been reporting high numbers of new cases in the first half of July, 2020, as high as 3.5K per day. As shown in Figure 5, the results from our model (calibrated until June 17) appear to be tracking the case surge quite well.

**Figure 5:**
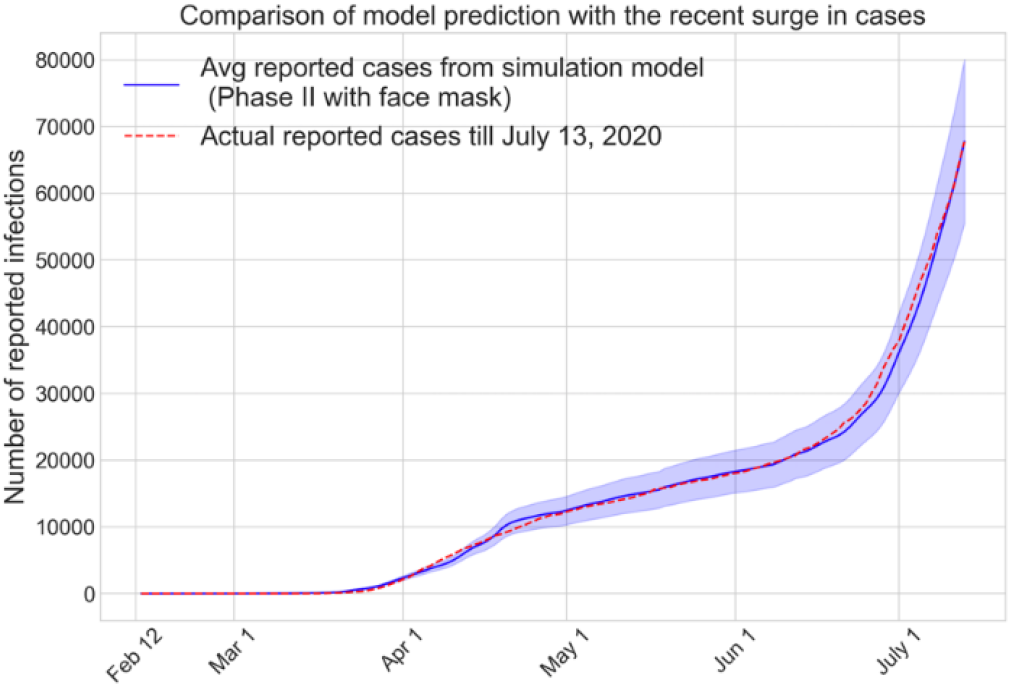
Simulation results track the surge in reported cases in July, 2020

The models used to predict COVID-19 outcomes so far have mostly been based either on observed data (e.g., [12-15]) or compartmental models like SEIR (susceptible, exposed, infected, and recovered) or their variants (e.g. [16-20]). Agent-based simulation models for COVID-19 can be found in [21, 22]. Observation data driven models are very well suited for understanding the past progression of a pandemic and also for estimating parameters characterizing virus epidemiology. However, data driven models offer limited prediction ability for the future especially in situations where the conditions on the ground (e.g., testing and treatment ability, social interventions, people’s behavior and response) change. SEIR type compartmental models guided by differential equations have been most widely used for communicable diseases, some early examples are [23-25]. Compartmental models are aggregate in nature and assume uniform behavior of the population over time. Hence, these models also do not adapt well to changing pace of disease transmission. An agent-based modeling approach is more suitable for a detailed consideration of individual attributes, specific disease natural history, and complex societal interventions [9].

Our agent-based model has several limitations. First and foremost, the simulation model is an abstraction of how a pandemic impacts a large and complex society. Though our model deliberately introduces some variabilities, somewhat pre-defined daily schedules are used to approximate a highly dynamic contact process of a urban region. Also, the contact process does not account for significant variabilities in the types and lengths of interactions even within each mixing groups. We did not assign geographic locations (latitude and longitude) for households, businesses, schools, and community places, and assumed them to be uniformly distributed over the region. It is common for urban population centers and associated facilities to grow in clusters, for which the contact patterns are different from those in dispersed regions. We did not consider special events like parties, games, and street protests, some of which is known to have caused superspreading of the virus and case increases. Finally, and perhaps most importantly, the model uses a large number of parameters (listed in Tables 1 through 11 in supplementary document) and hence the model predictions are influenced by the choice of those values. We have used published data from the government archives and research literature for most parameters. In absence of established data source, we have used expert opinion and media reports. The model results, as presented in this paper, are only expected outcomes based on currently available information.

Each scenario of our case study with 10 replicates (with different seeds) takes approximately 8-12 hours to run in a standard desktop computer with Intel Core i7 with 16GB memory. In the interest of presenting out observations quickly to the public health decision makers, while COVID-19 is still rampant in the region, we chose to use a limited number (10) of replicates. As the main purpose of this paper is to conduct a broad what-if analysis, we do not believe that use of a small number of replicates has negatively influenced our observations. The trends and observations derived from our results are intended to be used for planning and guidance of public health decision makers.

## Data Availability

All data used in the study are either from openly available government archives or from published research literature. All relevant references are provided in the data tables. Some of the information used are based on expert opinion and media reports.

## Author Contributions

Conceived and designed the model: TKD and HT; Selection of model input parameters and data gathering: RD, HT, and TKD; Coding and testing of the model: HT and TKD; Design and perform the experiments: HT and TKD. Output analysis and review: TKD, HT, and RD. Manuscript preparation and review: TKD, HT, and RD.

